# A qualitative evaluation of a multi-modal cancer prehabilitation programme for colorectal, head and neck and lung cancers patients

**DOI:** 10.1101/2022.11.01.22281795

**Authors:** Sharon Linsey Bingham, Sarah Small, Cherith Jane Semple

## Abstract

**Background:** Growing evidence indicates patients’ survivorship outcomes can be enhanced through active engagement in a multi-modal cancer prehabilitation programme (MCPP), although this intervention is not uniformly embedded as a standard of care. MCPP aims to optimise patients physiologically and psychologically for cancer treatments, shorten recovery time, reduce complications, promote healthier lifestyles and improve quality of life. South Eastern Health and Social Care Trust (SET) developed and evaluated a system-wide collaborative approach to MMCP across three tumour groups (colorectal, lung, head and neck cancer). Addressing the lack of qualitative evaluation of MCPPs, this paper explores mechanisms promoting feasibility and acceptability of MCPP from patients’ and interdisciplinary professionals’ perspectives.

**Methods:** Semi-structured virtual one-to-one interviews were conducted with 24 interdisciplinary professionals and nine patients. Transcripts were recorded, transcribed verbatim and themes developed using Framework Analysis.

**Results:** Analysis of findings identified four themes providing an in-depth understanding of key elements required to develop and deliver a MCPP: 1) Planning: Building the team, 2) Shared vision to develop and tailor the MCPP to meet patient needs, 3) Delivering the MCPP and 4) Moving forward - improving the MCPP.

**Conclusion:** The system-wide collaborative approach to developing a MCPP at SET was deemed both feasible and acceptable. Success was attributed to visionary leadership, alongside a diverse group of interdisciplinary professionals being engaged, motivated and committed to improving patient outcomes. Iterative, responsive troubleshooting during delivery facilitated successful implementation. Greater adherence to provision of prescriptive high intensity exercise within the programme may further promote enhanced patient outcomes.

**Key elements required to deliver a successful multimodal cancer prehabilitation programme**

- Necessity of visionary, strategic leadership
- Committed interdisciplinary stakeholders to include patient and public involvement
- Skilled, trained multi-disciplinary multi-sectoral team to deliver programme
- Screening of patients to tailor referrals to multimodal elements
- Effective and efficient referral processes
- Patient-centred pathways, delivered across universal, targeted and specialist stepped-care model based on patient’s needs
- Promote tailored intense exercise prescriptions
- Data collection parameters agreed and implemented to enable robust evaluation
- Effective and regular communication with professionals operationally delivering the programme
- Sustainable funding

## Background

Despite advancements in cancer treatment, 15–40% of patients with cancer who undergo surgical treatment experience postoperative complications [1]. As a result, patients may experience increased hospital stays or hospital readmissions, with negative impacts to physical functioning, psychosocial outcomes and overall quality of life [2]. Macmillan Cancer Support and the Royal College of Anaesthetists suggest that cancer prehabilitation should underpin the whole cancer pathway [3]. Taking forward this guidance, the Cancer Strategy for NI 2022-2032 [4] proposes that cancer prehabilitation should be available to all those who will benefit. Despite this recommendation and the growing body of evidence, cancer prehabilitation is currently not embedded as a standard of care within cancer care pathways locally, nationally or internationally [5]. This is due to a myriad of reasons, including workforce shortages, funding, lack of evidence around best practice models and need for addition information on cost-effectiveness [5-7].

Multi-modal cancer prehabilitation (MCPP) aims to optimise physical and psychological health through delivery of a series of tailored interventions including exercise, nutrition, and psychological support alongside behaviour change support for alcohol consumption and smoking [8], from the time of cancer diagnosis to the beginning of acute treatment. Most supporting evidence for MCPP is provided for the following tumour groups, colorectal (CRC), lung and head and neck (HN) [1,9-11]. Studies evaluating the efficacy of MCPP have discovered patient benefits even when implemented for just two weeks prior to treatment [12]. Patient benefits can include improved physiological function and resilience, shorter recovery time, reductions to peri-operative complications, gaining a sense of control over uncertainties ensued from a cancer diagnosis, improving quality of life and positive impacts on long-term health through behaviour change [12-13]. While people regain more control over their own health, the NHS and other healthcare systems have the potential to make economic savings [6].

A cancer unit within South Eastern Health and Social Care Trust (SET), United Kingdom, systematically planned, developed, implemented and evaluated a MCPP across CRC, lung and HN tumour groups during the COVID-19 pandemic with no additional funding [14]. The SET MCPP is a personalised and tailored MCPP, with referral pathways for individual’s based on screening using validated measures. The intervention focuses upon functional, emotional and nutritional needs aligned to patients’ tumour group and current lifestyle habits (smoking and alcohol) using the universal-targeted-specialised conceptual framework endorsed by Macmillan Cancer Support, the Royal College of Anaesthetists and National Institute for Health Research Cancer and Nutrition Collaboration [15] (see Fig 1). The universal and targeted exercise pathway aimed to have patients completing 3 high intensity training (HIIT) sessions per week, and the specialist pathway involved a physiotherapist-led bespoke cardiovascular and strengthening session per week with prescribed home exercise for alternate days. Universal emotional support was provided by Macmillan Move More Co-ordinators (MMCs) who had Level 4 Personal Training alongside generalist emotional support training; with targeted and specialist pathways delivered by counsellors /assistant or clinical psychologist respectively. Similarly, universal nutritional advice was provided by MMCs, with targeted and specialist input by dietetic support worker or dietitian. Smoking cessation support was provided when necessary, taking a motivational interviewing approach and providing advice on pharmacotherapies. Substance misuse liaison provided alcohol advice, education, and signposting to referral services for patient as indicated. The purpose of the MCPP was to improve patient function prior to treatment and optimise health and wellbeing [14]. From an organisational perspective the MCPP at SET sought to develop and harness collaborative interdisciplinary working across a range of partners, promote good practice, foster shared learning, inform restructuring of services, scale-up and roll-out of similar programmes. Given the complexity of SET’s MCPP [14] which comprised of interplay between interdisciplinary roles across a multi-sectoral team being planned and implemented in a real-world environment, qualitative methods were deemed appropriate for evaluation. This would enable an enhanced understanding of processes and mechanisms leading to MCPP’s success and failure, paramount prior to scaling up and further roll-out [16]. To date, the authors are aware of only one MCPP qualitative feasibility study, however the focus was solely upon delivery fidelity and acceptability [17]. This novel paper aims to explore mechanisms promoting feasibility and acceptability of a MCPP from patients’ and professionals’ perspectives exploring planning, development and implementation phases. The objectives were to gain understanding of mechanisms that:

**Fig 1.**
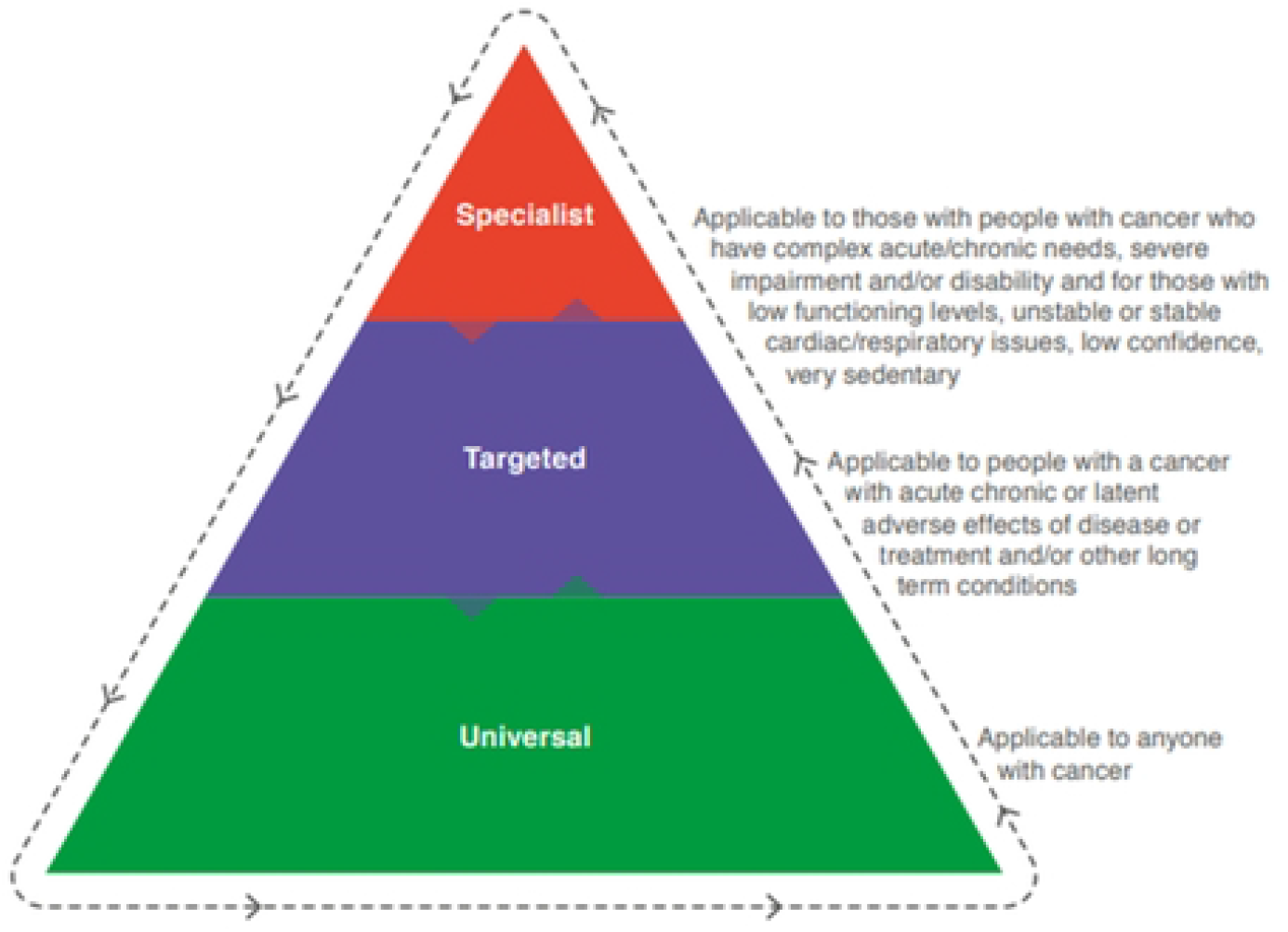
Stepped care framework for prehabilitation intervention [5].

1. promoted and informed planning and development of MCPP
2. promoted delivery of a personalised and tailored MCPP from patients’ and professionals’ perspectives
3. promoted acceptability and feasibility of MCPP implementation in a real-life context from patients’ and professionals’ perspectives.

## Method

### Study design

The overarching theoretical framework used was qualitative descriptive design [18], employing semi-structured interviews to explore interdisciplinary professional and patients’ perspectives of the MCPP.

### Sample and recruitment

A purposeful sample of 30 eligible interdisciplinary professionals across pivotal roles, involved in the planning, development, screening or delivery of MCPP was selected. See Table 1 for inclusion criteria. The professional sample included representation from medical, clinical nurse specialist (CNS), allied health professionals (AHPs), Move More Co-Ordinators (MMCs) and administration roles within SET, local councils and Macmillan Cancer Support. To ascertain interest in participation, potential interdisciplinary professionals received an email from the Principal Investigator (CJS), with study invite, participant information sheet and consent form. The second participant study population was patients, who were purposefully selected to maximise variation in terms of gender, engagement (drop-outs and completers) and tumour group (CRC=10, Lung=5, HN=5) and contacted via email or telephone by a CNS who acted as the Local Collaborator. These potential participants were also provided with a study invite, participant information sheet and consent form. Participants corresponded directly with SLB, a post-doctoral researcher external to SET, known to two professionals and unknown to all patients, to ask questions and confirm consent. All participants were recruited to the study between February and May 2022. Ethical approval for the study was granted by Ulster University Nursing and Health Research Filter Committee.

**Table 1.** Inclusion / Exclusion criteria for semi-structured interviews.

|  | **Inclusion criteria** | **Exclusion criteria** |
| --- | --- | --- |
| **Interdisciplinary professionals** | • Registered healthcare professionals (HPs) (CNS, AHP, medical staff), MMCs, health development and health informatic personnel directly involved in the development, screening, referral, or delivery of MCPP at SET<br>• Able to provide informed consent | • Those meeting inclusion criteria but on leave during data collection period<br>• Healthcare students |
| **Patients** | • Age >18 years old<br>• Diagnosis of CRC, lung or HN cancer<br>• Referred to SET MCPP<br>• Able to provide informed consent<br>• Communicate in English | • Patients deemed too unwell by relevant CNS at time of data collection<br>• Receiving end-of-life treatment<br>• Dementia or cognitive impairment |

### Data collection

To facilitate exploration of feasibility and acceptability, the Theoretical Framework of Acceptability (TFA) [19] and the Normalising Process Theory (NPT) [20] guided the development of topic guides used with each study population. The TFA is a multi-faceted framework to guide the assessment of acceptability based on anticipated, experiential, cognitive and emotional responses to the intervention, including constructs of affective attitude, burden, coherence, perceived effectiveness and self-efficacy. The NPT comprises of four components which seek to understand what individuals and groups do to enable an intervention to be normalised. These components are: 1) coherence (sense-making), 2) cognitive participation (engagement), 3) collective action (work to enable intervention to happen) and 4) reflexive monitoring (formal and informal appraisal of the benefits and costs of the intervention). Furthermore, topic guides developed were informed by literature and knowledge from subject experts and piloted prior to data collection. Semi-structured interviews were conducted by SLB using a virtual platform or telephone. Interviews were recorded and transcribed verbatim by either SLB or professional transcriber. Average interview duration was 28 minutes and 15 minutes for professionals and patients respectively. Quotes included in the evaluation had repeated words and unnecessary colloquial phrases e.g. “you know” removed, and the interviewer added detail within [] to improve coherence.

### Data Analysis

Transcripts were checked for validity (SLB) and read by all authors. Transcripts were coded by SLB in NVivo V12, with CJS independently coding four transcripts to enhance study rigour. Transcripts were analysed using Framework Analysis [21] and informed by the aforementioned theories (TFA and NPT). Framework analysis sits outside any epistemological perspective providing the researcher team flexibility in analysis, while providing a method to account for perspectives from a diverse group of participants [21]. Data from first 10 interviews were coded inductively and subsequent transcripts were deductively coded, in line with evaluation objectives. Initial inductive codes were developed and collated by the first and last author (SLB), and through team discussion initial themes identified. Through iterative team discussions (CJS, SS, SLB) the themes were refined, and final themes were established to ensure the correct meaning of the participants had been captured.

## Results

Participating interdisciplinary professionals (n=24) (n=6 no response) represented a range of disciplines, tumour groups, and sectors (see Table 2) and 9 patients represented 3 tumour groups with 11 declining participation as outlined in Table 3.

**Table 2.**
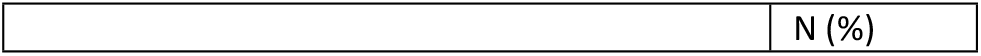
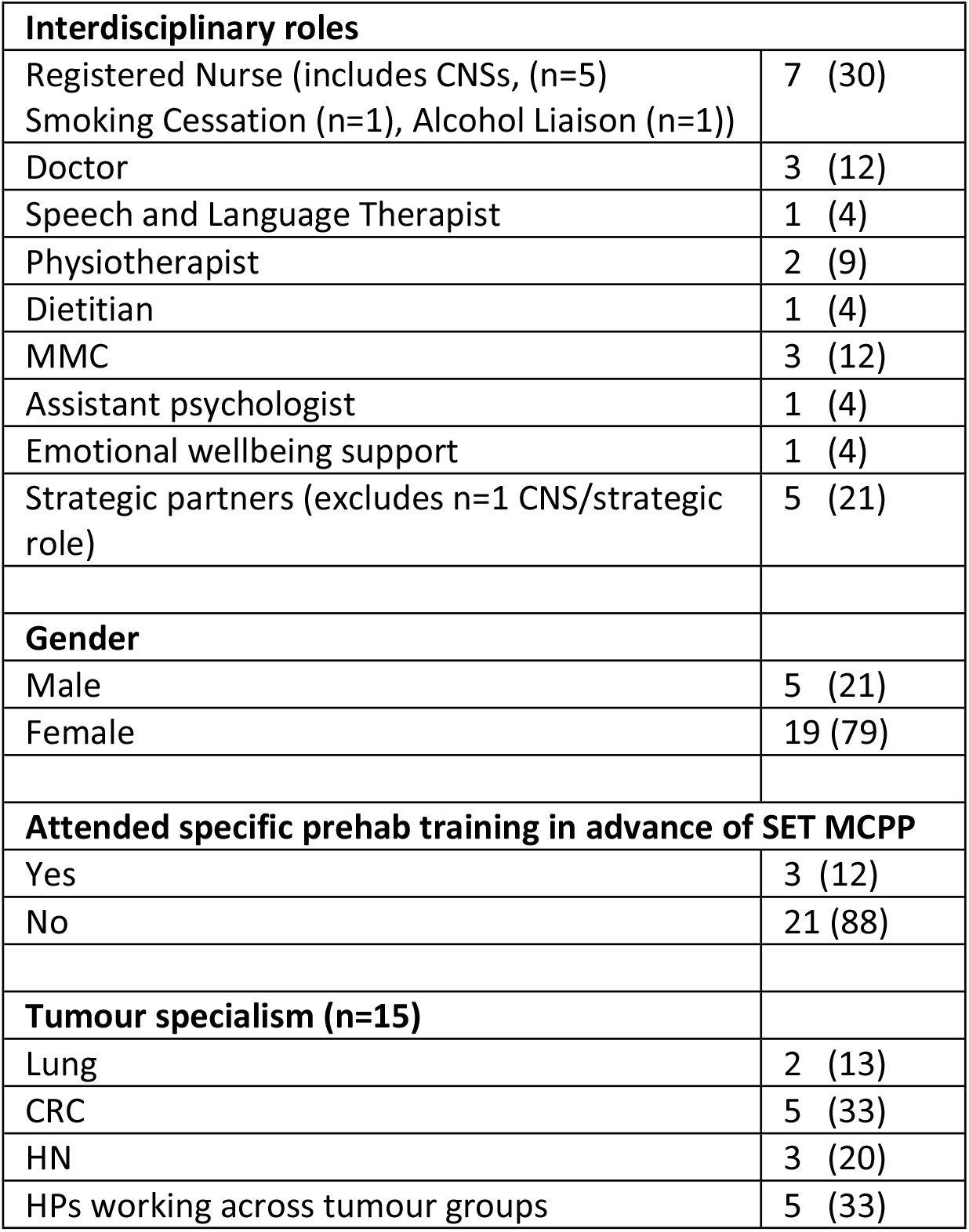
Demographics of interdisciplinary professionals.

**Table 3.** Patient demographics (n=9).

|  | N (%) |  | N (%) |
| --- | --- | --- | --- |
| **Engagement with evaluation (n=20 approached)** |  | **Support Network** |  |
| Accepted | 9 (45) | Very good | 8 (89) |
| Approached - No response | 6 (30) | Poor | 1 (11) |
| Initially interested – Withdrew, unknown reason | 3 (15) |  |  |
| Declined - No recollection of MCPP | 1 (5) | **Educational status** |  |
| Declined – Unwell | 1 (5) | Secondary school | 5 (56) |
|  |  | Third level education | 4 (44) |
| **Tumour group** |  |  |  |
| HNC | 2 (22) | **Frequency of exercise prior to diagnosis** |  |
| Lung | 4 (45) | Regular | 6 (67) |
| CRC | 3 (33) | Somewhat active | 2 (22) |
|  |  | Never | 1 (11) |
| **Age** |  |  |  |
| 46-60 | 2 (22) | **Access to video platform** |  |
| 61-75 | 6 (67) | Yes | 8 (89) |
| 76+ | 1 (11) | No | 1 (11) |
| **Gender** |  | **Comfort using video platform** |  |
| Male | 6 (67) | Very confident | 4 (44) |
| Female | 3 (33) | Fairly confident | 1 (11) |
|  |  | Not confident | 4 (44) |
| **Marital status** |  |  |  |
| Living alone | 1 (11) | **How was the MCPP delivered?** |  |
| Married/Living with partner | 7 (78) | Telephone | 3 (33) |
| Living with family member/ friend | 1 (11) | Virtual and telephone | 3 (33) |
|  |  | In-person | 4 (44) |
| **Time from Diagnosis** |  |  |  |
| 1-2 months | 1 (11) |  |  |
| 6-12 months | 7 (78) |  |  |
| 12 months + | 1 (11) |  |  |

Four themes were identified from professional and patient data, 1) Planning: Building the team, 2) A shared vision that led to successful development of MCPP: the important elements, 3) Delivering the MCPP: in action and 4) Moving forward - improving the MCPP.

### 3.1 Theme 1: Planning: Building the team

Having clearly established the purpose of the team, namely to plan, develop, deliver and evaluate a MCPP at SET there was a need to build a diverse interdisciplinary team to enable delivery of the tailored MCPP across the agreed stepped-care conceptual framework. Attributable characteristics for professionals was enthusiasm and motivation, with strategic and clinical champion roles. The vision and energy for programme delivery was clear from the outset.

> *“It’s great that we’re able to pilot it here in the SE Trust … there was a study in Manchester, … the research and the data that’s come out of that, its proven to be a really, good way of getting patients back up on their feet. So, it’s brilliant.” P17 (Emotional support)*

Key to the successful engagement of HPs was their previous knowledge of Enhanced Recovery After Surgery and the growing evidence-base for cancer prehabilitation. Furthermore, what appeared important for HP engagement was team leadership with credible expert clinical experience and inspiring and motivating leadership style. P18 (Smoking Cessation) stated,

> *“To be fair, [Strategic clinical programme lead] can motivate people and get people around the table and do that’s kind of exactly what made such a difference.”*

Collective, positive ‘can do’ attitudes towards the MCPP pilot programme were key to exploring the potential of and progressing the MCPP within current resources during COVID-19, with P22 (Pelvic Physiotherapist) reporting,

> *“I sort of went in, you know, very open minded, I really wasn’t really sure what this was going to entail.”*

Overwhelmingly professionals viewed MCPP as a worthwhile intervention with reports of how MCPP was “a “no-brainer” (P20 strategic). Anticipated benefits and motivating factors for professionals was the desire that MCPP could improve patient outcomes, alongside formalising pathways for care delivery and reduced costs. P1 (CNS HNC) highlighted MCPP as an

> “*Opportunity to give some control back to the patient … and is a prime opportunity to enable and engage people early in a shared care model of their treatment*.”

Professionals’ knowledge and prior experience of MCPP varied; some with more limited knowledge, while others were well-informed through conference attendance and abreast of published research. MMCs, while qualified in their field, lacked experience supporting patients at diagnosis. P2 (MMC) reported,

> *“Traditionally, with Move More, we would have seen those [patients] sort of post-treatment, probably in treatment and post-treatment, post-treatment would have been our biggest*.”

This lack of experience did impact upon confidence when engaging with patients early in the treatment trajectory, identifying a need for training and practice supervision.

### 3.2 Theme 2: A shared vision that led to successful development of MCPP: the important elements

Reporting on the early development of the MCPP through to implementation, this theme encompassed three subthemes which highlight elements related to [1] Developing the MCPP, [2] Person-centredness and [3] Attention to professionals’ training needs.

#### [1] Developing the MCPP

With a shared vision, the strategic steering group organised five timebound (3 weeks) Task and Finish groups incorporating views of subject matter experts on exercise, nutrition, emotional support, data management and evaluation. Assisted by experts in Performance Management an electronic referral process was developed which along with screening tools, pathways and outcome measures was “*all explained in the Standard Operating Procedure”* [P24 MMC]. Furthermore, contributing to a foundation of trust and formalising governance structures, data sharing agreements were established between SET and local council partner organisations along with secure channels for sharing patient information.

Interdisciplinary professionals collaborated to select robust, validated, appropriate and easy to use screening and assessment tools to inform the patient’s tailored referral pathway and enable evaluation of the MCPP. Selection was achieved through a consensus approach with the team seeking to strike a balance between *“the usability and the time burden and practicality element of things.”* (P5 Strategic). Further adaptations to outcome measures were required in response to COVID-19 restrictions; the 6-minute walk test was substituted for a 30 sec sit-to-stand test with similar efficacy to facilitate remote assessment.

#### [2] Person-centred approach to delivery

The screening process enabled a personalised and tailored prehabilitation programme for patients, making the MCPP a vital component of the cancer care continuum. HN cancer services piloted a ‘One-stop Prehab clinic’ whereby patients, following diagnosis and on completion of staging scans, had a scheduled appointment to meet with a CNS and AHPs including SLT and dietetics in central dedicated clinical space, reducing appointments for patients and enhancing communication regarding patient need between specialities. The CNS, dietetic and SLT perceived advantages to this approach, as patients could move easily between HPs and HPs could ensure co-ordinated provision of informed care from diagnosis throughout treatment trajectory. P19(SLT) described,

> *“We had a clinic a Monday afternoon … usually, CNS saw them first, and then speech therapy next, and then dieticians … logistically, it was a good way of working … we had the benefit of CNS … [who] could give us any updated information…so it was really helpful.”*

Unfortunately, the SLT reported that due to workforce pressures their discipline is not currently represented at this ‘One-stop Prehab clinic’ for patients with HN cancer but indicated a keen willingness and desire to re-establish this clinical commitment when adequate funding was available to support service delivery.

#### [3] Attention to professionals’ training needs

Many of the HPs reported feeling adequately equipped for their role in the MCPP with transferrable skills, with some finding informal direction on specific issues as sufficient. For others, development of the Standard Operating Procedures manual helped to equip them to deliver the MCPP. Some AHPs who did not routinely work within cancer care, reported their delivery of a multi-modal element within the MCPP was more challenging, recommending a need for specific training for team members from a non-cancer background. More often, this was gaining knowledge on best practice when communicating with patients who have received a potentially life-limiting diagnosis, whilst for others this was related to disease-specific clinical information.

> “*I really didn’t have very much knowledge about even the surgery that is involved with cancers, so a big learning curve, really, for myself*.” (P22 Pelvic Physiotherapist)

MMC’s received formal training from Macmillan Cancer Support on “Managing difficult conversations” and “Motivational Interviewing” (P24 MMC). Furthermore, MMCs were also able to access training provided by SET subject experts covering areas like ‘Healthy eating – nutrition’, ‘Smoking cessation’, ‘Alcohol and You’, exercises to promote “*core stability*” for patients with CRC cancers and “*head, neck and shoulder exercises*” for patients with HN cancers (P1 CNS HNC). MMCs were also provided with training relating to collecting data using functional and patient-reported outcome measures. When issues hindering implementation of the MCPP across disciplines were identified, further appropriate and timely training was offered to enhance fidelity of the intervention in accordance with the ‘Standard Operating Procedure’ manual. Training was flexible and aligned to need of professional, with personnel providing training considered as “*extremely approachable and contactable*” (P21 CNS HNC).

### 3.3 ***Delivering the MCPP: in action*** (within this theme are patients and professionals experiences of the MCPP)

Selection of patients for MCPP was based on predetermined inclusion and exclusion criteria, which included tumour staging and a range of physiological parameters. However, it would appear that some CNSs across the three tumour groups, were selecting patients who they perceived would benefit most and considered to be motivated and engaged, raising the possibility of HPs acting as gatekeepers to the programme.

HPs (mainly CNS and consultants) staged communication with patients and family members regarding MCPP, with initial conversations succinctly introducing the topic, presenting a brief outline of programme components and benefits with their written patient resource pack. Subsequent conversations with other professionals delivering elements of the programme provided more detail to reduce information burden at diagnosis.

> *“I think probably at diagnosis the patient can be overwhelmed with what has been told. But [we tell them], we’re sending home the resource pack and saying, ‘Look don’t focus too much about that, just at the moment, you will get a phone call, and it’ll be all explained better at that time.’”* (P21 HNC CNS)

Most patients offered MCPP were interested although HP presence may have added bias. Patients regarded MCPP as a credible approach with Patient 7 describing the MCPP as “*it’s a no brainer*,” a descriptive phase similarly used by professionals. Patients believed that participation in the MCPP would enhance physical preparedness for surgery, aid post-treatment recovery and inform future care. Patients described reassurance, comfort and connectedness to the healthcare team knowing there would be support from the outset of their cancer trajectory.

*“When you get your diagnosis of cancer, you sit there and you get this diagnosis that you spend your entire life, hoping you never do*… *you just think your world has ended, you know, and being reached out to by somebody who is saying, no. You know, we’re going to we can stay positive through this. We can stay moving, we can keep in touch, you can stay moving… And I found that even that alone, the reach out, somebody’s reaching out and saying, no, no, come on, XXXX [Patient name] we’re going to do this.”* (Patient 1 Lung)

Patients who did opt in to the MCPP, were reported by HPs to engage well and thankful for the specialist input early on in their treatment journey. When there was resistance, HPs perceived this to be due to patients feeling overwhelmed, the patient’s perceived lack of need for such an intervention and resistance to behaviour change.

Screening tools and the online referral form were largely viewed positively however the Distress Thermometer was identified as problematic given the baseline distress at cancer diagnosis, but its inclusion agreed important as described by P4 (CNS CRC).

> *“The Distress Thermometer was barely an accurate one, because they [patients] are so extremely anxious…so if it is ridiculously high … they need referred for the Associate Psychologist, you know, for that other than that universal emotional pathway, so I think it’s by no means an accurate measurement but I suppose it still has to be a baseline.”*

Electronic screening and the online referral process employed for SET MCPP were viewed favourably. Completion of one online form per patient, enabling referral to different pathways within the conceptual multimodal stepped-care framework, was deemed highly acceptable and advantageous. This approach was keenly driven to avoid “*the copious referrals sometimes necessitated when assessing new patients*” (P1 CNS HNC Strategic) whilst enabling a tailored MCPP approach. P1 (CNS HNC Strategic) describes this as,

> “*A referral form … to just tick and select the components with the prehabilitation and personalise that and with one submit button automatic referrals would generate to the relevant people, so that was my vision so thankfully we’ve been able to action that which is one of the great wins in the system that we do have today*”

This online form was easy to complete and use by CNSs, although technical issues were identified. That said, early implementation was hampered by incomplete forms with P21 (CRC CNS) reporting *“I don’t think I realised the importance of filling in every single detail*,” although rectified by prompts.

Delivery of MCPP was enhanced by well-structured and action orientation meetings, which supported effective team working and created an environment of mutual respect and shared learning. Meetings facilitated interdisciplinary working, enhanced understanding of clinical and non-clinical roles, interdisciplinary learning and real-time troubleshoot.

> *“My work [is] not cancer focused. I was meeting other professionals that I’ve never ever had been dealing with and involvement with Health Development [Team] …also the Councils and Macmillan … so that I just felt it was fabulous, the way everybody worked together… I just felt that the working together and the troubleshooting and the problem solving was great.”* (P14 Physiotherapist)

Information received through electronic referral was reported as largely sufficient, however, when necessary, patient’s Electronic Care Records (ECR) were accessed and reported as a useful bolster. For some HPs, they had a desire for clearer information on patient’s treatment and prognosis to inform care.

> *“I think sometimes that’s maybe a bit unfair on the patient of us turning up, you know, because if somebody is basically going to be dead in two months, I mean, is it really appropriate to start going on at them about their alcohol use if it’s been a big crutch in their life.”* (P6 Alcohol Liaison)

The Alcohol Liaison and Smoking Cessation Services, reported processes were akin to already developed approaches within HNC, however patient interest in engaging with these services varied, as P18 pointed out,

> *“Obviously, if somebody is going through a cancer diagnosis, that they can be anxious, they can be agitated. And that’s always reasons for them to carry on smoking. So, it’s a bit of a challenge.”* P18 (Smoking Cessation)

Nonetheless, referrals to the MCPP were considered as appropriate with most professionals having capacity within their role to deliver MCPP. MMCs placed most emphasis on the exercise component seeking to ensure that exercise prescription fitted with patient’s individual need, capability, motivation and routine. However, at a strategic level there was some concern that the exercise prescription by MMC was being viewed “*in its loosest form*” (P12 MMC), with reduced credence on HIIT. Effort to address this was initiated through further booster training session with MMCs. This appeared to have limited impact with MMCs believing interventions like HIIT to be unrealistic for a number of patients referred to the MCPP. On the other hand, patients viewed the exercise programme positively; reflecting on its varied, patient-centred and achievable attributes, including indoor and outdoor options which Patient 6 (Lung) reporting that “*it helped me get out of the house too*.” MMCs were trained to provide nutritional advice and emotional support under universal pathway; however, findings highlighted that MMCs focused less on these elements attributed to perceptions that this was not the MMC’s main area of expertise.

> *“I wouldn’t say it’s [nutritional advice provision] very in depth on any level, you know what we’re not nutritionally trained and I suppose like any physical activity coordinator, somebody comes looking nutritional in-depth advice, you know, we refer them on because that’s not our forte, we’re not trained in that, but we do have basic knowledge and understanding.”* (P2 MMC)

Patients demonstrated positive perceptions of the MCPP, being equipped with adequate information, highlighting the information pack was comprehensive and allowed them to digest the information at home. Furthermore, patients reported feeling informed about their treatment, having an increased confidence because of the MCPP support, and emotionally supported at what was a difficult time as described by Patient 4 (Lung) *“[If] you were having a bad day, just being in their company would give you a boost.”*

Patients also highlighted benefits to fitness, reducing or quitting smoking and alcohol, and a sense their treatment outcomes benefited as a result, Patient 6 (Lung) highlighted their biggest benefit,

> *“Well, it was it was the exercises that the nurse was doing with me….You know all in all it seemed dead simple…You can get help whenever you want. There’s just so simple, but the difference it was unbelievable… he [Surgeon] said my fitness was fine to do surgery.”*

Of note, family members supported and saw value in the MCPP, with the reflection that patients received additional support during the pandemic when they were often isolated. Patient 8 (HNC) reported,

> *“I was talking to my, my daughter in law, who, unfortunately has terminal cancer and has been living with us for a few years now…she said it was nice, that I was able to talk to these people [MCPP professionals], have the support and know exactly what was ahead … she didn’t have that facility*.

There was a great sense of patient gratitude for the MCPP, with patients impressed by professionals’ commitment and availability to communicate with patients between appointments. Patient 4 (Lung) commented,

> *“I think it’s something that will benefit patients and the Health Service both. I thought it was well-managed, well-run, and just [want to] say thank you.”*

### 3.4 Moving forward – improving the MCPP

This theme will be presented as three subthemes as professionals and patients reported a series of [1] barriers and [2] facilitators linked to the implementation of the MCPP at SET and made several suggested important [3] improvements moving forward.

#### Subtheme [1]: Barriers to implementing MCPP

Overarching challenges to MCPP implementation related to the timing of the intervention, its delivery timeframe and the impact of COVID-19 restrictions on in-person contact. MCPP was most often introduced at diagnosis, while necessary to maximise prehabilitation timeframe, HPs reported that patients were often feeling overwhelmed with little capacity to process further information. The often-short timeframe between diagnosis and treatment was challenging for MMCs who reported that patients had to sort personal affairs; but delivery was further impacted by COVID-19 guidance on isolation. P5 (Strategic) described,

> *“We’ve had to adapt with the [COVID-19] guidance, you know, the patients are obviously being swabbed and at different stages, you know, so if you think about a pathway, a patient’s been screened, they’ve been referred, they might have the time one [data collection], but then they’re getting swabbed and told right your operations next week, but you have to isolate for 72 hours before your operation, you know, so it’s adjustments within the COVID guidance at the time to keep them on a green pathway for their treatment.”*

Spurious theatre lists during COVID-19 often meant that patients were provided with limited notice for date of surgery, with MMCs having less time than anticipated for MCPP and inability to capture Time 2 functional and patient-reported outcome measures (post-MCPP and before commencement of cancer treatment). Striking a balance between data collection and intervention delivery was often difficult, with MMCs advocating for a longer programme delivery timeframe (at least 2-weeks) to ensure optimal data collection and delivery of multimodal components.

Virtual communication approaches used for working group meetings, on one hand facilitated attendance at meetings, but for some inhibited interaction. MMCs were somewhat resistant to virtual approaches to service delivery, although recognised virtual platforms as a safer method to telephone contact when collecting functional data collection, particularly for the 30-second Sit-to Stand test. Variation in COVID-19 restrictions for local councils meant delivery mode of MCPP differed across the four local council areas, reflective of varied MMCs’ employment and governance structures. For some MMCs there was no in-person visits during the pandemic due to their local council guidance, for others, garden visits were permitted, making not only the development of a therapeutic relationship easier, but enhanced collection of functional measures such as Grip Test. Rapport building for the delivery of targeted emotional support was also thought to be hampered by virtual communication methods, as P10 (Assistant Psychologist) reported,

> *“She [the patient] mentioned a few times how frustrated she was with not being able to meet somebody face to face. You know, have that part, that connection, I suppose.”*

However, in-person contact was not welcomed by all patients as P5 (Strategic) described,

> *“Patients are being told to isolate, they’re worried about it jeopardising their surgical treatment, you know, they didn’t want to go to leisure centres or see people they don’t need to see*.” (P5 Strategic)

Despite these professional reported challenges, most patients did not experience barriers to the MCPP due to mode of intervention delivery. Although cognisant of COVID-19 restrictions, more often patients indicated a preference for in-person contact, and when unavailable one patient highlighted his unease with online platforms. Patient 3 (HNC) described,

> *“I’m not a fan of video and zoom calls. So, if I could have got either a telephone conversation or face to face, I was much happier*.”

#### Subtheme [2]: Facilitators for the MCPP

The collaborative effort by a team of interdisciplinary, multi-sectoral enthusiastic and committed professionals facilitated the implementation of MCPP in SET. Key drivers to its successful implementation were visionary leadership, shared vision and commitment of professionals throughout the MCPP planning, development and implementation phases, which enabled MCPP to become increasingly embedded into an integrated pathway. Effective communication between skilled, committed professionals and patients contributed to MCPP success. Patient 4 reported,

> *“[LUNG CNS] and the team did encourage me but not in a pressured way. It was very much voluntary. I didn’t feel obliged. My family were very supportive. So that all seemed to be entirely sensible, and a good thing to do.”*

As MCPP processes were increasingly embedded into HPs’ practice, the perceived time burden for screening and referral by the CNSs was reduced, as P4 (CNS CRC) reflected,

> *“I guess it was just really with practice, that we got into the habit of getting all the correct information. So, it did take a wee while initially, a lot of information wasn’t being teased out.”*

In addition, timely and interval communication on dissemination outputs and sharing outcome-based accountability reporting cards kept interdisciplinary professionals abreast of progress and facilitated ongoing commitment.

COVID-19 restrictions, while creating barriers provided unique opportunities. For example, the Health Development team had unexpected time capacity to opportunistically develop elements of MCPP at SET, which resulted in better co-ordination across tumour groups and services.

> *“So, I think for us to have gone ahead and gone on our own. It might have worked, it might not, but it would have been much more difficult. This is a much, much better model.”* (P13 Physiotherapist)

Furthermore, changes to service delivery as a result of COVID-19 has left a legacy of a hybrid MCPP delivery model, deemed as helpful by some patients, in creating flexibility and reducing travel time to appointments. Also, inclusion of family members or partners was perceived to support patient engagement, particularly when patients relied on them for transport to appointments. P12 (MMC) described,

> *“So, you know, maybe somebody’s saying, ‘Oh, I don’t drive, but my husband might be able to bring me in’, [and I would reply] well sure, he can come and take part in or he can do this, or he can do that.”*

Some professionals reported skill development as a consequence of being engaged in the process, such as working with patients with cancer, project management, collaborative multisectoral working, promoting job satisfaction from contributing to positive patient outcomes. Those involved in the MCPP reported an increased sense of value in their work, with satisfaction sharing project progress with other healthcare Trusts and Department of Health. P8 (Dietetic) reflected,

> *“I think you definitely feel like it’s another sort of element to the dietetic role that it’s because you are part of the MDT [multidisciplinary team] and part of the project, it’s a lot more, you feel very valued I suppose and your input very beneficial.”*

#### Subtheme [3]: Improving MCPP

To aid an enhanced and sustained MPCC at SET there are several key priorities areas worth noting. Recurrent funding was the most pressing reported priority, thus enabling a sustained model of funding for interdisciplinary roles to take forward MCPP as P3 (Strategic) reported *“I think sometimes we underestimated the amount of time and expertise required to really shape and drive the project forward”*. Funding is required to create equitable services across trust boundaries, as geographical limitations to services prevented patients referred from other Health and Social Care Trusts (HSCTs) to elements of the MCPP. Funding should include provision for project management, professionals’ roles including MMCs, AHP specialities, data management personnel and account for project scale-up (incorporating further tumour groups), while taking account for staff leave.

Effective communication and service level agreements between professional groups were also considered as paramount, aiding clarity to reporting structures particularly when working across organisations. Due attention is required when introducing new personnel into established yet novel systems, necessitating careful induction. P10 (Assistant Psychologist) reported, “*So, I didn’t even know it existed….I didn’t even know what cancer prehab was.”* Regular attendance at meetings was key with P1 (CNS HNC) describing,

> “*Having those continued and sustained on a regular basis and for me is never make assumptions that those delivering a service are actually delivering*.” (P1 CNS HNC)

Furthermore, timely communication with MMCs relating to patient treatment schedules could improve collection of Time 2 data and intervention tailoring. A review of the data collection timepoints and a renewed consensus agreement on achievable expectations may prove helpful and improve data capture.

Flexible training with booster sessions was welcomed and is required on an ongoing basis to promote intervention fidelity and equip new personnel. To enhance this, developing role competencies for MCPP could help to identify training needs. Consequently, this would outline expectations for MMCs regarding universal nutritional advice and importance of exercise prescription, potentially promoting greater patient benefits. P5 (Strategic) identified that,

> *“[MMCs] need to be prescriptive … patients do need to be pushed outside their comfort zone … a lot of it [current provision of MCPP] is self-directed, …they’re [patients] still kept in their comfort zone of what they’re happy to do, you know, walking twice a week, or whatever it is.”* (P5 Strategic)

Lastly, the development of a bespoke IT system, which is patient-centric to enable seamless screening, referral and assessment would, while costly, be advantageous by reducing reliance on Performance Management role co-ordinating data collection across three assessment points, reducing professional burden and system error.

For the patient, MCPP could be improved through feedback mechanisms when using remote delivery of universal elements of the services, reassuring patients that exercises were undertaken correctly. Finally, the opportunity to hear about a previous patient’s experience was regarded as a beneficial when used in other parts of care, with patients reporting it as “*one of the best things that happened*” and “*very reassuring*” when they felt “*scared stiff*” (Patient 3 HNC). Patients recommended that this should be adopted as part of the MCPP.

## 4. Discussion

This qualitative evaluation provided an enhanced understanding of mechanisms leading to the feasibility and acceptability of the MCPP in SET; in conjunction with important attributes that could be changed or adopted to augment scaling up of the intervention. Despite the COVID pandemic backdrop, interdisciplinary collaboration across multisectoral organisations forged ahead without funding, maximising upon innovation and creativity to integrate pathways for a MCPP, striving for better cancer patient-outcomes at SET. Success was mainly due to the visionary leadership and diverse and committed group of interdisciplinary professionals working together to adapt and refine a tailored stepped-care MCPP for local implementation at a Cancer Unit in Northern Ireland [14]. Current research investigating the feasibility and acceptability of MCPPs from the perspectives of professionals and patients is exceptionally limited [17] with studies often referring to patient recruitment and participation as indicators of success [23-24]. The findings from this study extend current evidence by illustrating the mechanisms of success and challenges experienced by those at SET and will be discussed through the lens of NPT components [20], coherence, cognitive participation, collective action and reflexive monitoring.

High levels of *<u>coherence</u>* were evident across interdisciplinary groups. Professionals were clear on how MCPP could complement and be a vital part of the cancer care continuum. Key facilitators, such as having knowledge that MCPP can positively impact patients’ functional and emotional outcomes, along with reducing the economic burden of patient care upon the healthcare budget [5,25-26] has promoted successful implementation of MCPP. Of note, for some, greater coherence could be facilitated through an enhanced appraisal of the recent growing body of literature surrounding the efficacy of MCPP. For example, some doctors in this study were less aware of the multifaceted components of the programme and known benefits, even within a short timeframe of less than two weeks [27]. Rectifying this is important as patients, both within this study and elsewhere articulate that doctors’ communication on the topic influences their uptake and engagement with MCPP [22]. High coherence can enhance patient recruitment and adherence [28]. Patients who understood the purpose and benefits of the programme were more committed to the programme, with the converse also evident. High coherence in the short time-period between cancer diagnosis and treatment commencing can be challenging to achieve given levels of diagnosis-related stress patients experience at the time of introduction to the MCPP, thus an evident need for professionals to be skilled in communication, facilitation and providing emotional support [29].

The MCPP at SET successfully engaged a varied and committed skilled workforce across tumour groups including those with strategic, clinical and physical activity knowledge and skills, reflecting good *<u>cognitive participation</u>* [19]. Partnering, through established links, with local councils and community and voluntary sector was advantageous as there was already established a sense of trustworthiness. Similar partnership arrangements were also reported as a key to success when Greater Manchester adopted a system-wide approach to MCPP [30]. Involvement from key professionals early in programme development and in an on-going manner was crucial to success, alongside ensuring that decision-making occurs in a collaborative and shared manner [22]. Cross-sectoral collaborations, however, can be challenging given differing reporting and funding structures [31]. This was somewhat the case for MMCs during the MCPP, which could be addressed by closer participative working with local council employers. One notable perspective less visible during the development phase of the MCPP was that of the patient. Best practice [32-33] states it is essential to involve end-users of the intervention throughout development to ensure that the intervention is acceptable, engaging and feasible, anticipating the needs of others, as often this not effective. Representative and proactive patient involvement may effectively address some of the difficulties with coherence mentioned above [25].

*<u>Collective action</u>* is characterised by how people work together to achieve outcomes, build confidence and organisational support [34]. Collaboration between engaged and envisioned professionals was integral to the development, implementation and ultimately the success of this programme. Each discipline had clear roles throughout development and implementation, reporting back on progress at working group meetings. Active participation throughout development and implementation contributed to a sense of self-efficacy, with professionals highlighting personal progress working with patients with a cancer diagnosis, screening and outcome tools. Complex interventions delivered in complex systems [35] require high intervention fidelity to accurately determine the impact of the intervention under study [36]. Delivery of the SET MCPP exercise component was critiqued by some HPs at a strategic level as requiring more specific and intensive exercise prescription, despite the agreed exercise doses outlined in the MCPP Standard Operating Procedure manual. Murdoch *et al*. [17] study of their MCPP with a CRC patient group suggested that while professionals emphasised that fitness was important for surgery outcomes, patients were not encouraged to increase activity levels. Herein, they noted that the lack of exercise prescription was attributed to a short timeframe between diagnosis and treatment, plus constrained resources with clinicians being too busy [17]. In contrast, the SET MCPP universal and targeted exercise prescription was delivered by MMCs within the local council setting who had capacity to deliver the programme components, yet exercise prescription was still considered to be lacking HIIT.

Future MCPP outcomes may benefit from additional training, monitoring of intervention delivery, and ensuring professionals integrate skills taught to enhance intervention fidelity [37]. It would be important that any quantitative evaluation focusing on objective functional and patient-reported outcome measures from the SET MCPP are understood in terms of the current perceived fidelity to the programme components.

*<u>Reflexive monitoring</u>* was evident throughout the SET MCPP with monthly working group meetings providing a vehicle to review progress and facilitate troubleshooting. Analysis, real-time feedback and booster training sessions allowed many issues to be addressed, however, some outcome data capture remained problematic. Given the often-short timeframe for MCPP delivery, alongside lack of knowledge of, or changeable, treatment commencement dates presented as challenges for MMCs to collect Time 2 data. Minimising attrition rates of Time 2 data capture is paramount to enhance the evidence base for MCPP [26]. Development and implementation of the MCPP at SET received no additional funding, however a sustained funding model is required to further consolidate, scale-up and integrate pathways for system-wide implementation. Professionals in this study, like that of others [26,38] recognised the need for dedicated funding which extends to sustaining AHP involvement [26], project management and performance data support [38]. Given the success of MCPP already evident [28], there is now a need to align funding to ensure equitable access of MCPPs to all cancer patients. The NPT components [20] has been useful to illuminate the mechanisms of feasibility and acceptability alongside key areas for refinement of the MCPP to maximise future implementation and to identify important considerations for others seeking to embark on implementation of a MCPP.

### Strengths & limitations

This study benefited from an exploration of a diverse range of patient and professionals’ perspectives. However, the Local Collaborations (CNSs) were potentially more likely to enrol patients considered as engagers onto MCPP and therefore this patient study sample may not be representative of a generic cancer population. During recruitment to this qualitative evaluation effort was directed in trying to engage patients who dropped out of MCPP, but this proved challenging depicted in the patient recruitment rate of less than 50%. It is possible that patients participating in this study are likely to have a positive bias towards MCPP. Further investigation is required to determine the barriers to participation.

### Conclusions

The MCPP at SET was deemed a feasible and acceptable tailored and personalised intervention; with visionary leadership, engaged and motivated interdisciplinary professionals, committed to improved patient outcomes being central to its success. Iterative, responsive troubleshooting throughout implementation contributed to embedding individualised elements of a MCPP. The MCPP could benefit from greater prescription of HIIT to maximise potential patient outcomes. Improved professional and patient knowledge of the benefits of MPCC could maximise referrals to MCPP and enhance patient outcomes.

## Data Availability

Data is available upon reasonable request from the primary author but not available publicly to promote anonymity of qualitative data collected from participants of one healthcare trust.

